# Estimating county-level flu vaccination in the United States

**DOI:** 10.1101/2023.05.10.23289756

**Authors:** Andrew Tiu, Shweta Bansal

**Affiliations:** Department of Biology, Georgetown University, Washington, DC, USA

## Abstract

In the United States, influenza vaccines are an important part of public health efforts to blunt the effects of seasonal influenza epidemics. This in turn emphasizes the importance of understanding the spatial distribution of influenza vaccination coverage. Despite this, high quality data at a fine spatial scale and spanning a multitude of recent flu seasons are not readily available. To address this gap, we develop county-level counts of vaccination across five recent, consecutive flu seasons and fit a series of regression models to these data that account for bias. We find that the spatial distribution of our bias-corrected vaccination coverage estimates is generally consistent from season to season, with the highest coverage in the Northeast and Midwest but is spatially heterogeneous within states. We also observe a negative relationship between a county’s vaccination coverage and social vulnerability. Our findings stress the importance of quantifying flu vaccination coverage at a fine spatial scale, as relying on state or region-level estimates misses key heterogeneities.

## Introduction

Seasonal influenza (flu) epidemics in the United States cause significant disease burden each year and are a recurring public health challenge. The Centers for Disease Control and Prevention (CDC) typically estimate flu-related illnesses in the tens of millions and flu-related hospitalizations in the hundreds of thousands each year [1]. Fortunately, seasonal flu vaccines are effective in reducing disease burden [2] and the Advisory Committee on Immunization Practices (ACIP) consistently recommends seasonal flu vaccines as safe and effective for all those six months and older [3], with some exceptions. Vaccination is especially important for certain groups, for example those 65 years and older, that are typically at higher risk for developing severe disease outcomes.

Despite this, our understanding of flu vaccination coverage in the US has significant gaps. While the CDC compiles survey responses pertaining to flu vaccination from sources like the Behavioral Risk Factor Surveillance System (BRFSS) surveys and the National Immunization Surveys (NIS)–collectively distributed as their FluVaxView data [4]–these are primarily at the state level, obscuring important heterogeneity at a finer scale. County-level versions of these vaccination data are available but are model estimates rather than data natively collected at the county level and are limited to only three recent flu seasons. And, although the Selected Metropolitan/Micropolitan Area Risk Trends of BRFSS [5] (SMART BRFSS) subset of the data contains some county-level information, it only focuses on a small collection (typically 4% each year) of counties in qualifying metropolitan and micropolitan statistical areas, meaning the spatial extent and utility of the data are limited. Additionally, while past studies have investigated historical US flu vaccination trends, especially among healthcare workers or older, more vulnerable populations [6–12], to our knowledge, there have been no national-level analyses that examined vaccination coverage in the broader population at a fine spatial scale and spanned recent seasons (namely 2020 onward).

Underlining the importance of understanding US flu vaccination patterns is the link between low vaccination and social vulnerability: the most vulnerable geographic regions typically have poor vaccination coverage. Past work has shown that social determinants of health like socioeconomic status, race, and urbanicity add an additional layer of complexity, influencing access to vaccines and manifesting as inequities in vaccination among the most vulnerable [13–15]. The CDC attempts to describe social vulnerability through its Social Vulnerability Index (SVI) [16], and prior studies have used this index to investigate and uncover vaccination-related disparities in the US [17, 18]. The SVI quantifies a community’s vulnerability to disasters, including natural disasters or disease outbreaks, based on a set of 16 social factors that are grouped into four themes.

Here, we estimate county-level flu vaccination in the United States for five consecutive flu seasons from 2018 to 2021. We leverage a massive commercial medical insurance claims data source to gather counts of flu vaccination while accounting for sample representativeness via survey raking techniques. To handle bias in these claims data, we use publicly-available, survey-based flu vaccination data to provide a state-based measure of bias. Finally, we explicitly incorporate these bias measures and the SVI in a set of Bayesian generalized linear models to produce more accurate county-level flu vaccination estimates than is currently available. Our model estimates illustrate significant geographic heterogeneity in influenza vaccine uptake, highlight low coverage in the most vulnerable communities, and emphasize the importance of monitoring vaccination coverage at a fine spatial scale.

## Methods

### Data source and processing

To characterize the landscape of seasonal flu vaccination in the United States, we use large-scale commercial medical insurance claims data. For analysis, we create a curated subset of this data source that contains claims information down to the county level for approximately 140 million individuals from 2016 onward. We can access individuals’ age and sex information and are able to filter their claims for specific diseases or medical services based on the presence of International Statistical Classification of Diseases and Related Health Problems (ICD) codes [19], Healthcare Common procedure Coding System (HCPCS) [20] codes, and National Drug Codes (NDC) [21]. Furthermore, this data source provides third-party estimates of certain demographic information like race/ethnicity.

To address sample representativeness issues in the insurance claims data, we use a survey raking approach via the `anesrake` [22] R package. Survey raking, or iterative proportional fitting, is a process by which individuals in a sample are assigned weights such that the sample distribution of a chosen set of (raking) variables matches the known population distribution [23, 24]. A benefit of this approach is that only the marginal population distributions are needed. For each year from 2016 to 2021 and for a given county, we perform raking on age (binned), sex, and race, using county-level 2021 US Census Bureau data [25] for the target distributions. Thus, each individual in a given county is assigned a year-specific weight. Because the raking algorithm using age, sex, and race may fail to converge, or some levels of a raking variable are not observed in the data set (making raking impossible), we also rake on age and sex only. These weights act as a fallback and may be used as a substitute for the aforementioned cases in which age-sex-race raking was not successful. This ensures every individual is assigned a weight and henceforth, we refer to this combined set of raking weights as “sub” weights.

We identify cases of flu vaccination by using a set of flu vaccine-related Current Procedural Terminology (CPT) [20] codes and 11 digit NDCs (Supplementary Table 1). We define county-level counts of flu vaccination by aggregating the number of individuals in a given county with a claim containing at least one flu vaccine-related CPT or NDC code during a given flu season. These seasons span July 1 of the first year to May 31 of the second year, and we collect data from the 2018-2019 flu season to the 2020-2021 flu season. To give a sense of vaccination coverage, we also obtain county-level counts of individuals with any recorded claim (all cause) during the corresponding flu season. Lastly, we use the “sub” raking weights to generate weighted versions of both of these counts and all further analyses are conducted with these weighted counts (Fig 1A).

**Figure 1:**
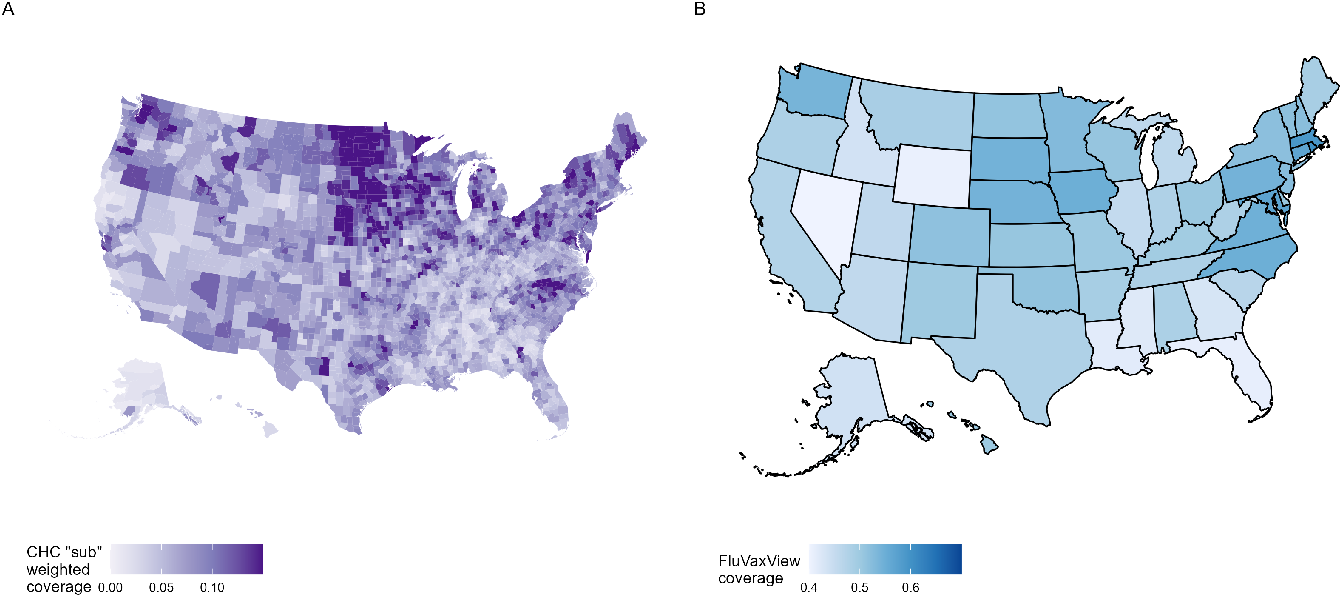
A selection of county-level flu vaccination coverage using raking weights (A) and statelevel flu vaccination coverage from the CDC’s FluVaxView data set (B), United States 2018-2019 flu season. A) Raking weights are a combination of weights generated by raking on age, sex, and race substituted for weights from raking on only age and sex, hence the label “sub” weights. Areas in the Northeast/East and Midwest show the highest coverage, with some pockets of higher coverage in the Pacific Northwest. B) FluVaxView coverage estimates roughly match the national pattern in vaccination, with regions on the east coast and in the Midwest having the highest levels of coverage, but neglect nuance found in finer-scale coverage.

### Bias estimate

To further combat sampling bias, namely the innate differences in sampling effort and coverage of the insurance claims data, we treat the state-level FluVaxView data provided by the CDC as ground truth. The FluVaxView dataset combines information from the NIS and BRFSS to cover all age demographics (Fig 1B). This process allows us to define a degree of bias for flu vaccination coverage derived from the insurance claims data. Specifically, we quantify bias as

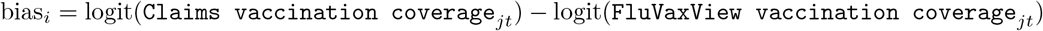

where *i* indexes a bias observation for state *j* in flu season *t*. We then use a simple frequentist hierarchical model (fit using the `lme4` R package) to take advantage of partial-pooling and obtain a single estimate of bias for each state:

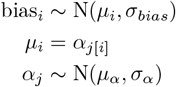

### Binomial regression model

To estimate the bias-corrected probability of flu vaccination in county *i*, we fit the following Bayesian aggregated binomial model for each flu season, where *M*_*i*_ is the number of individuals vaccinated in county *i, N*_*i*_ is the total number of patients in county *i* (*M*_*i*_ ≤ *N*_*i*_), *p*_*i*_ is the probability of vaccination in county *i*, and *x*_*i*_ is the SVI (2018 overall theme) value for county *i*. Index values range from zero (lowest vulnerability) to one (highest vulnerability) and represent a percentile ranking. We include the model estimates of bias as an offset term for county *i* in state *j*:

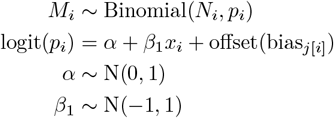

The models are fit using the `brms` [26, 27] R package with `cmdstanr` [28] backend. We specify the sampler to use four chains and 4000 iterations per chain. The models successfully converge, with all 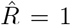 and all parameters’ bulk-effective sample size (bulk-ESS) well over 1500, indicating sufficient mixing of the chains and strong estimation accuracy.

Finally, the bias-corrected vaccination coverage *v*_*i*_ is calculated as the following, where *η*_*i*_ is the posterior linear predictor:

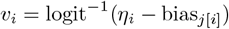

## Results

To characterize county-level flu vaccination in the US, we collect counts of seasonal flu vaccination harnessing a rich data set of medical insurance claims. After applying raking to generate a more representative sample of vaccination counts, we use aggregated binomial regression to model the probability of flu vaccination in each flu season, controlling for potential bias in the data by explicitly quantifying bias relative to robust third-party survey data collected at the coarser, state level.

### Predicted patterns of county-level flu vaccination in the United States are highly heterogeneous but vary little across seasons

Bias-corrected predictions of flu vaccination coverage at the county level show similar spatial patterns from season to season. Across all seasons, predicted coverage ranges from 42.2% to 56.4%, with the mean in each season typically lying just above 50%. Our models consistently predict the highest levels of vaccination coverage in counties in the Midwest/West and Northeast (Figure 2), especially for seasons prior to the 2020-2021 season. Counties in the South/Southeast and West Coast tend to have the lowest coverage, though there exist small pockets of high coverage in these regions as well. We also find a negative association between vaccination coverage and SVI, with more vulnerable counties having lower vaccination levels.

**Figure 2:**
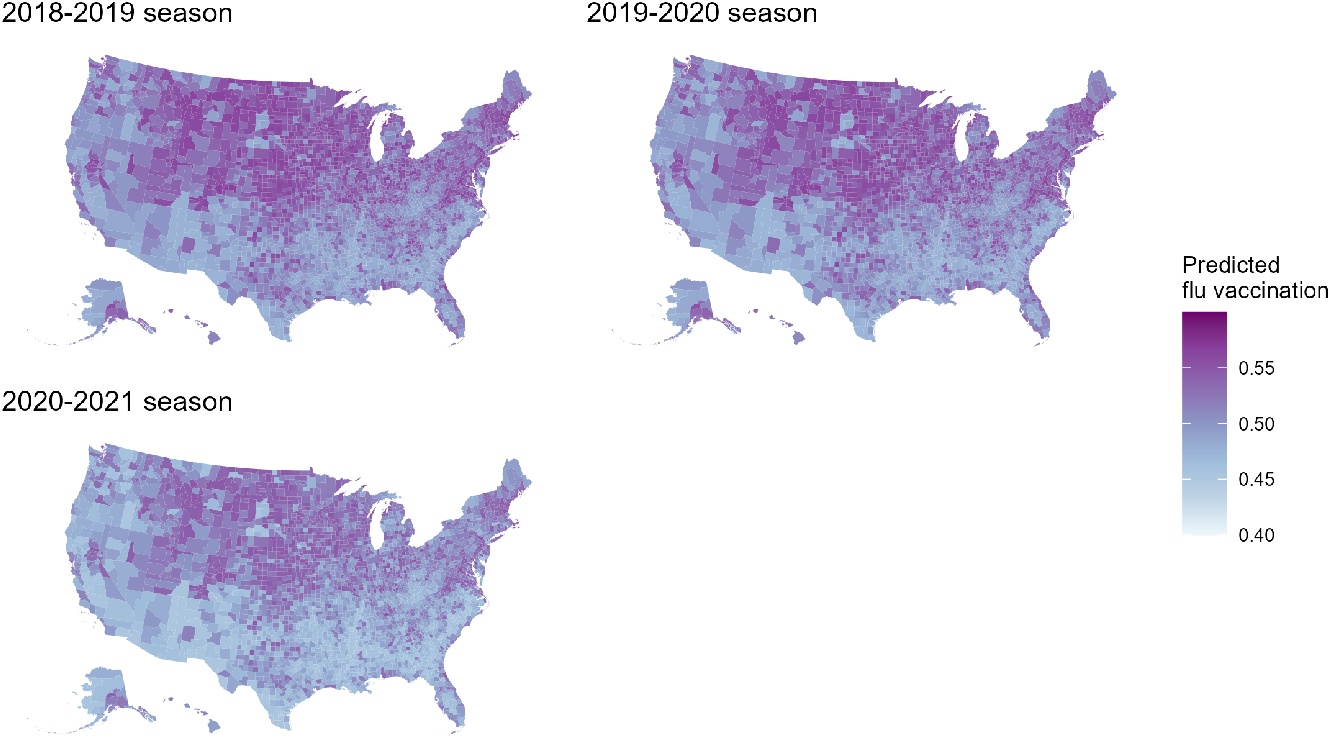
Bias-corrected predictions of flu vaccination coverage for seasons from 2018 to 2021. Points are colored by state. Predictions are highest in the Midwest/West and Northeast. This spatial pattern appears steady across flu seasons.

Our bias-corrected predictions match well with corresponding survey-derived FluVaxView estimates. In the majority of seasons the state means of county-level predictions lie close to the identity line when compared with state estimates (Figure 3. However, county-level vaccination still varies within a state and is not simply dictated by the state-level coverage.

**Figure 3:**
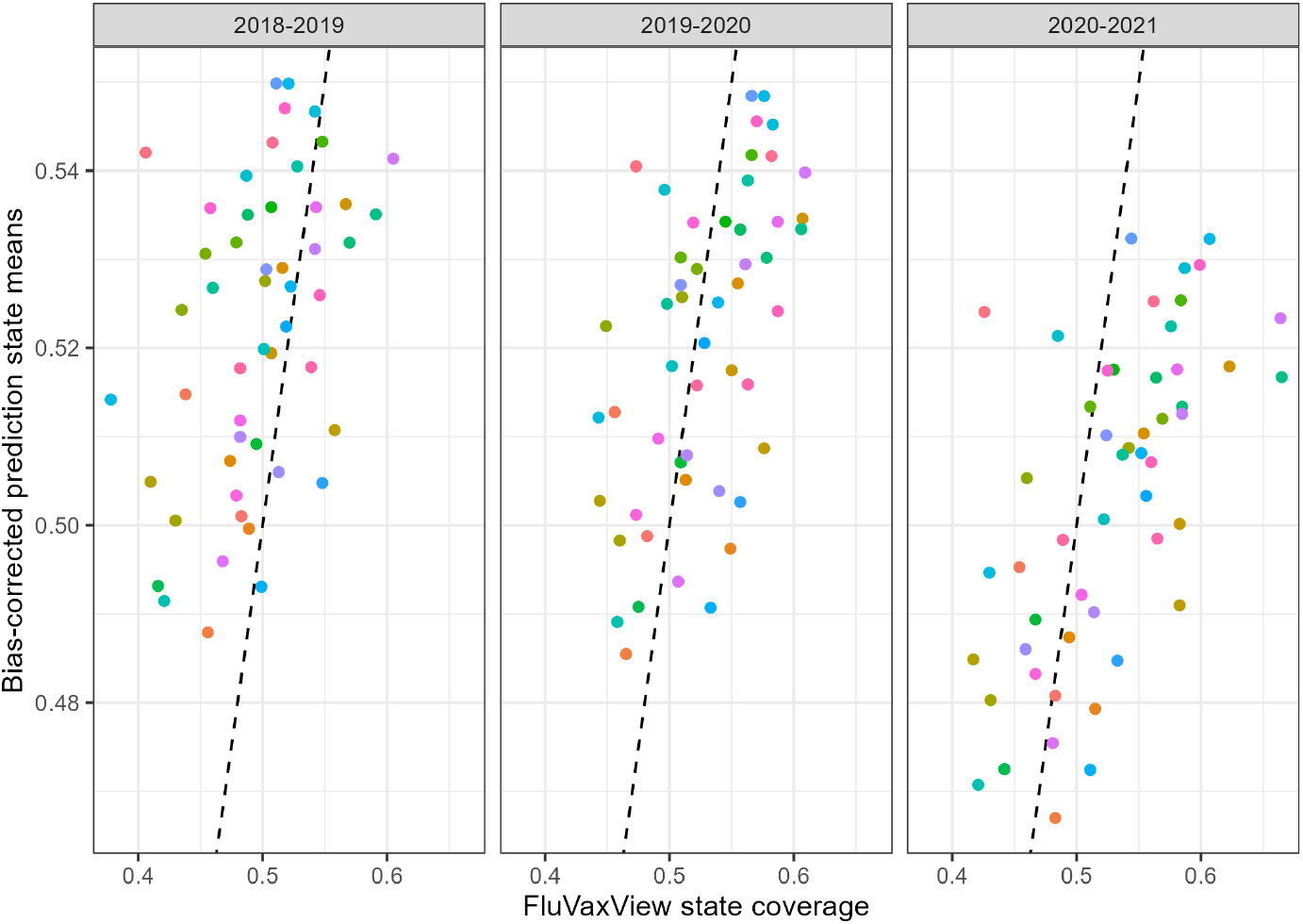
Comparison of state means of bias-corrected predictions with FluVaxView state coverage levels. County-level predictions for each state are averaged, with colors representing each state, and the identity line (black, dashed) also shown. State means of bias-corrected predictions generally match FluVaxView state coverage.

## Discussion

Although flu vaccination plays a key role in public health plans to tackle seasonal influenza epidemics in the US, high quality, county-level estimates of vaccination are not readily available for a wide range of seasons. The CDC’s FluVaxView data set integrates vaccination information from a multitude of carefully designed surveys and represents a gold standard for assessing coverage levels [4]. However, these data are largely limited to state-level estimates, and county-level estimates are only available for three seasons, with these finer-scale estimates being model-generated predictions rather than purely based on survey responses. Additionally, while SMART BRFSS data are technically available at the county level, these are limited to a subset of locations based on the number of survey responses from that area [5]. This lack of fine-scale flu vaccination data hinders efforts to understand coverage patterns, accurately identify areas of undervaccination, and design public health interventions.

Our modeling approach takes advantage of a large set of medical insurance claims to address this gap in data. The insurance claims data have the advantage of being at the county-level and, along with demographic information, allow us to weight vaccination counts to obtain a more representative sample of counts. By combining these weighted counts with aforementioned Flu-Vax View estimates, we take advantage of this survey-based information to explicitly incorporate bias from these counts and then make bias-corrected predictions that are at the county level. We find that patterns of flu vaccination are largely consistent between seasons. Areas in the southern/southeastern parts of the US and along the West Coast continuously have the lowest predicted flu vaccination coverage. These southern/southeastern regions are typically the most vulnerable parts of the country from a public health standpoint [15, 29] and often experience the start of the flu season the earliest [30, 31], deepening the need to address undervaccination and better tailor public health efforts here. Furthermore, this pattern of flu vaccination coverage generally aligns with observations for other vaccines or vaccination in general [32–34], and this consistency likely points to wider issues surrounding healthcare inequality and/or poor healthcare infrastructure in these communities.

Accurate, consistent, and fine-scale characterization of flu vaccination patterns should be a key part of public health plans. However, such efforts should not be solely limited to the flu vaccine. As we demonstrate here, there is significant utility that clearly extends to other types of vaccines. Besides collecting vaccination data at a finer scale like the county level, there are benefits to using other existing, diverse data sets and simple modeling frameworks to achieve more accurate views of vaccination across the country.

## Data Availability

All data produced in the present study are available upon reasonable request to the authors.

## Acknowledgments

Author affiliations: Department of Biology, Georgetown University, Washington, District of Columbia, United States.

AT collected the data, performed all analyses, interpreted results, and drafted the manuscript. SB designed the study, guided analysis, and edited the manuscript. All authors have read and approved the final manuscript.

This work was supported by the National Institute of General Medical Sciences of the National Institutes of Health under Award Number R01GM123007.

The authors declare no competing interests.

## Supplementary Information

**Table 1:** NDC and CPT codes for detecting claims related to seasonal flu vaccination.

| Code type | Codes |
| --- | --- |
| NDC | 33332-0110-10, 33332-0010-01, 33332-0113-10, 33332-0113-11, 33332-0013-01,33332-0013-02,33332-0114-10,33332-0114-11,33332-0014-01,33332-0014-02,33332-0015-01,33332-0015-02,33332-0115-10,33332-0115-11,33332-0116-10,33332-0116-11,33332-0016-01,33332-0016-02,33332-0117-10,33332-0117-11,33332-0017-01,33332-0017-02,33332-0118-10,33332-0118-11,33332-0018-01,33332-0018-02,33332-0319-01,33332-0319-02,33332-0419-10,33332-0419-11,33332-0219-20,33332-0219-21,33332-0220-20,33332-0220-21,33332-0420-10,33332-0420-11,33332-0320-01,33332-0320-02,33332-0421-10,33332-0421-11,33332-0221-20,33332-0221-21,33332-0321-01,33332-0321-02,33332-0416-10,33332-0416-11,33332-0316-01,33332-0316-02,33332-0317-01,33332-0317-02,33332-0417-10,33332-0417-11,33332-0318-01,33332-0318-02,33332-0418-10,33332-0418-11,66521-0000-01,66521-0000-11,70461-0001-01,70461-0001-11,70461-0002-01,70461-0002-1,70461-0018-03,70461-0018-04,70461-0019-03,70461-0019-04,70461-0020-03,70461-0020-04,70461-0120-03,70461-0120-04,70461-0121-03,70461-0121-04,58160-0879-52,58160-0879-41,58160-0880-52,58160-0880-41,58160-0881-52,58160-0881-41,58160-0883-52,58160-0883-41,58160-0900-52,58160-0900-41,58160-0901-52,58160-0901-41,58160-0903-52,58160-0903-41,58160-0905-52,58160-0905-41,58160-0907-52,58160-0907-41,58160-0898-52,58160-0898-41,58160-0896-52,58160-0896-41,58160-0885-52,58160-0885-41,58160-0887-52,58160-0887-41,42874-0012-10,42874-0012-01,42874-0013-10,42874-0013-01,42874-0014-10,42874-0014-01,42874-0015-10,42874-0015-01,42874-0016-10,42874-0016-01,42874-0017-10,42874-0017-01,42874-0117-10,42874-0117-01,49281-0718-10,49281-0718-88,49281-0719-10,49281-0719-88,49281-0720-10,49281-0720-88,49281-0721-10,49281-0721-88,63851-0612-01,63851-0612-11,62577-0613-01,62577-0613-11,62577-0614-01,62577-0614-11,63851-0613-01,63851-0613-11,70461-0200-01,70461-0200-11,70461-0419-10,70461-0419-11,70461-0319-03,70461-0319-04,70461-0420-10,70461-0420-11,70461-0320-03,70461-0320-04,70461-0321-03,70461-0321-04,70461-0421-10,70461-0421-11,70461-0301-10,70461-0301-12,70461-0418-10,70461-0418-11,70461-0201-01,70461-0201-11,70461-0318-03,70461-0318-04,19515-0889-07,19515-0889-02,19515-0890-07,19515-0890-02,19515-0893-07,19515-0893-02,19515-0850-52,19515-0850-41,19515-0845-11,19515-0845-01,19515-0895-11,19515-0895-01,19515-0894-52,19515-0894-41,19515-0891-11,19515-0891-01,19515-0898-11,19515-0898-01,19515-0901-52,19515-0901-41,19515-0903-11,19515-0903-01,19515-0908-52,19515-0908-41,19515-0912-52,19515-0912-41,19515-0896-11,19515-0896-01,19515-0909-52,19515-0909-41,19515-0900-11,19515-0900-01,19515-0906-52,19515-0906-41,19515-0897-11,19515-0897-01,19515-0816-52,19515-0816-41,19515-0818-52,19515-0818-41,66019-0110-10,66019-0110-01,66019-0107-01,66019-0108-10,66019-0108-01,66019-0109-10,66019-0109-01,66019-0300-10,66019-0300-01,66019-0302-10,66019-0302-01,66019-0301-10,66019-0301-01,66019-0303-10,66019-0303-01,66019-0304-10,66019-0304-01,66019-0305-10,66019-0305-01,66019-0306-10,66019-0306-01,66019-0307-10,66019-0307-01,66019-0308-10,66019-0308-01,66521-0112-02,66521-0112-10,66521-0116-10,66521-0116-11,66521-0116-02,66521-0116-12,66521-0117-10,66521-0117-11,66521-0117-02,66521-0117-12, |
| NDC | 66521-0118-10,66521-0118-11,66521-0118-02,66521-0118-12,70461-0119-10,70461-0119-11,70461-0119-02,70461-0119-12,70461-0120-10,70461-0120-11,70461-0120-02,70461-0120-12,66521-0113-10,66521-0113-02,66521-0114-02,66521-0114-10,66521-0115-02,66521-0115-10,49281-0386-15,49281-0010-10,49281-0010-50,49281-0387-65,49281-0010-25,54868-6177-00,54868-6180-00,49281-0388-15,49281-0011-10,49281-0111-25,49281-0011-50,49281-0012-10,49281-0012-50,49281-0705-55,49281-0112-25,49281-0390-15,49281-0392-15,49281-0392-78,49281-0707-55,49281-0707-48,49281-0113-25,49281-0113-00,49281-0013-10,49281-0013-58,49281-0013-50,49281-0013-88,49281-0014-50,49281-0014-88,49281-0394-15,49281-0394-78,49281-0396-15,49281-0396-78,49281-0391-65,49281-0393-65,49281-0393-88,49281-0395-65,49281-0395-88,49281-0397-65,49281-0397-88,49281-0399-65,49281-0399-88,49281-0401-65,49281-0401-88,49281-0403-65,49281-0403-88,49281-0405-65,49281-0405-88,49281-0120-65,49281-0120-88,49281-0121-65,49281-0121-88,49281-0389-65,49281-0709-55,49281-0709-48,49281-0703-55,49281-0708-40,49281-0708-48,49281-0710-40,49281-0710-48,49281-0712-40,49281-0712-48,49281-0413-50,49281-0413-88,49281-0513-25,49281-0513-00,49281-0413-10,49281-0413-58,49281-0621-15,49281-0621-78,49281-0414-50,49281-0414-88,49281-0414-10,49281-0414-58,49281-0514-25,49281-0514-00,49281-0415-10,49281-0415-58,49281-0516-25,49281-0516-00,49281-0416-50,49281-0416-88,49281-0416-10,49281-0416-58,49281-0625-15,49281-0625-78,49281-0627-15,49281-0627-78,49281-0417-50,49281-0417-88,49281-0517-25,49281-0517-00,49281-0417-10,49281-0417-58,49281-0518-25,49281-0518-00,49281-0629-15,49281-0629-78,49281-0418-50,49281-0418-88,49281-0418-10,49281-0418-58,49281-0519-25,49281-0519-00,49281-0419-10,49281-0419-58,49281-0419-50,49281-0419-88,49281-0631-15,49281-0631-78,49281-0420-50,49281-0420-88,49281-0633-15,49281-0633-78,49281-0420-10,49281-0420-58,49281-0520-25,49281-0520-00,49281-0421-10,49281-0421-58,49281-0421-50,49281-0421-88,49281-0521-25,49281-0521-00,49281-0635-15,49281-0635-78,58160-0808-15,58160-0802-02,58160-0804-01,76420-0482-01,76420-0483-0 |
| CPT | 90630, 90653, 90654, 90655, 90656, 90657, 90658, 90660, 90661, 90662, 90664, 90666, 90667, 90668, 90672, 90673, 90674, 90682, 90685, 90686, 90687, 90688, 90689, 90694, 90756, G0008, Q2034, Q2035, Q2036 Q2037, Q2038, Q2039 |

